# Assessment of the Feasibility of Remote Training, At-Home Testing, and Test-Retest Variability of Clustered Virtual Reality Perimetry

**DOI:** 10.1101/2022.10.07.22280753

**Authors:** Zer Keen Chia, Alan W. Kong, Marcus L. Turner, Murtaza Saifee, Bertil E. Damato, Benjamin T. Backus, James J. Blaha, Joel S. Schuman, Michael Deiner, Yvonne Ou

## Abstract

**Objective:** To assess the feasibility of remotely training glaucoma patients to take a ten-session clustered virtual reality (VR) visual field test (VVP-10) at home, analyze results for test-retest variability, and assess correspondence with conventional perimetry.

**Design:** Cross-sectional study.

**Subjects:** 21 subjects with glaucoma were enrolled and included in the feasibility assessment of remote training. 36 eyes were used for test-retest analysis and determination of concordance with Humphrey Visual Field (HVF) testing.

**Methods:** Subjects were provided with a mobile VR headset containing the VVP-10 test software and trained remotely via video conferencing. Subjects were instructed to complete ten sessions over a 14-day period.

**Main Outcome Measures:** Feasibility was determined by the number of subjects who were able to independently complete VVP-10 over the 14-day period after one remote training session. Intraclass correlation coefficient (ICC) of average fraction seen across ten sessions and standard error (SE) for the mean were primary outcome measures for assessing test-retest variability. Correlation with HVF mean sensitivity (MS) across eyes, was a secondary outcome measure.

**Results:** 20 subjects (95%) successfully completed the VVP-10 test series after one training session. ICC of VVP-10 was 0.95 (95% CI [0.92, 0.97]). Mean SE in units of fraction seen was 0.012. The Spearman correlations of VVP-10 average fraction seen versus HVF MS were 0.88 (95% CI [0.66, 0.99]) for moderate to advanced glaucoma eyes, and decreased to 0.68 (95% CI [0.29, 0.94]) when all eyes were included.

**Conclusions:** Remote training of patients at home is feasible and subsequent remote clustered visual field testing using VVP-10 by patients on their own without any further interactions with caregivers or study staff was possible. At-home VVP-10 results demonstrated low test-retest variability. Future studies must be conducted to determine if VVP-10, taken at home as convenient for the patient, may be a viable supplement to provide equivalent or complementary results to that of standard in-clinic assessment of visual function in glaucoma.

## Introduction

Visual function is routinely evaluated in glaucoma patients with conventional static automated perimetry (SAP), for example with Humphrey Visual Field (HVF) testing. However, patients dislike taking the test and report feeling pressured to do well.^1^ Additional challenges include variable testing environments, long clinic wait times, and the need to schedule appointments and travel to clinic.^1^ Furthermore, visual field tests have high inherent variability due to their short duration which limits the amount of data collected. Variability is also influenced by factors including the testing environment and technician instructions,^2,3^ and the need for patients to maintain prolonged fixation throughout the test can cause fixation losses, fatigue, and discomfort.^1,4^ Current tests also require significant clinic infrastructure and resources.

Studies have demonstrated that time to detection of progression using standard visual field testing regimens can range from 2 to 7 years or more depending on frequency of testing and rate of progression of disease,^5^ with factors such as ethnicity and socioeconomic status affecting the frequency of visual field testing,^6^ time to detection of progression,^7,8^ and severity of disease at first presentation.^9^ Chauhan et al. and the European Glaucoma Society therefore recommend performing six visual field examinations within the first 2 years of diagnosis to identify rapid progressors and establish patients’ baseline data,^10,11^ but a study by Fung et al. revealed that most patients with newly-diagnosed glaucoma received less than 3 visual fields in the first 2 years following diagnosis.^12^ Furthermore, while the American Academy of Ophthalmology recommends at least yearly visual field evaluations in patients with primary open-angle glaucoma,^13^ Stagg et al. found that more than 75% of patients with open-angle glaucoma across the United States receive less than one visual field test per year and thus do not meet guideline-adherent monitoring.^14^ Given the challenge of obtaining consistent visual field tests, especially during the COVID-19 pandemic, there is an unmet need for accessible, patient-friendly visual field testing that would not only enhance the patient experience but also provide physicians with additional data to help deliver the best care possible. One logical alternative to in-clinic visual field testing is at-home testing which patients are amenable to because it enables closer monitoring of their visual function through clustered testing and allows them to undergo testing at their convenience.^15,16^

Vivid Vision Perimetry (VVP, Vivid Vision Inc., San Francisco, CA) has designed a variety of portable and mobile virtual reality-based visual field tests that are configured to run on off-the-shelf head mounted displays using the principle of multi-fixation, ^17,18^ multiple-choice perimetry. The patient’s task during VVP was designed to reduce fatigue as compared to the button-press task in conventional in-clinic perimetry, with the aim of collecting more data and thus more precise measurements of sensitivity in patients with moderate to severe loss. Briefly, with VVP testing, patients are not required to fixate on a stationary central target for the duration of the test but are instead allowed to move their eyes and head during the test. A fixation task is used to control the retinal positions of the stimuli, although VVP is not a form of eye-movement perimetry and eye position is not monitored. Visual field testing is one of the least popular tests done in clinic,^4^ possibly in part due to the need for patients to suppress their foveation reflex, and multi-fixation perimetry may provide a more comfortable testing experience for patients. Indeed, patients report that tests that do not require constant fixation are more comfortable than SAP.^15,19^

In this study, we introduce VVP-10, a test with suprathreshold stimuli comprising ten identical sessions that patients complete over a 14-day period at home after remote training. The previously reported suprathreshold screening version of the test, VVP Swift, demonstrated good test-retest reliability, but only two tests per subject were included in the study.^17^ The precision of patients’ visual field measurements is limited both by the number of tests they receive each year and by the number of stimuli presented per test, so we sought to increase precision by increasing the number of sessions per patient and by using a new version of the test that presents stimuli more often at each location.

The purpose of this study was to demonstrate the feasibility of remote training of glaucoma patients to complete VVP-10 at home, to assess their at-home test results for test-retest variability using measurements obtained from the series of sessions, and to assess patient discomfort and fatigue. A secondary aim was to determine concordance between the visual field results obtained from VVP-10 and from HVF.

## Methods

Participants for this cross-sectional study were recruited from the patient cohort of the Glaucoma Clinic at the University of California, San Francisco (UCSF), from May 2020 to August 2020. Informed consent was obtained from all participants either in clinic or over Zoom (San Jose, CA). All methods were approved by the UCSF Institutional Review Board, and all research adhered to the tenets of the Declaration of Helsinki and the Health Insurance Portability and Accountability Act.

### Subject Characteristics

Inclusion criteria included subjects aged 18-85 with a diagnosis of primary open-angle glaucoma or secondary forms of glaucoma including steroid-induced, mixed-mechanism, and pseudoexfoliation glaucoma. A diagnosis of glaucoma was made by either optic disc or retinal nerve fiber layer (RNFL) structural abnormalities, reliable and reproducible visual field abnormalities consistent with RNFL damage defined as persistent scotomas on at least 2 consecutive HVF tests, or both. Structural abnormalities included neuroretinal rim thinning, localized or diffuse RNFL defects, disc hemorrhages, or progressive narrowing of the neuroretinal rim with increased cupping, observed with slit-lamp biomicroscopy and a handheld lens or with spectral-domain OCT imaging (Optovue Inc., Fremont, CA). Subjects who met our inclusion criteria were identified prior to each clinic day, and they were recruited in the order of their visits until our team ran out of Oculus devices; we did not seek to recruit subjects who were younger or otherwise potentially more technologically literate.

Exclusion criteria included best-corrected visual acuity (BCVA) worse than 20/80 or concurrent retinal disease including retinal vein occlusion, wet age-related macular degeneration, or proliferative diabetic retinopathy. Subjects with a history of epilepsy or issues with neck strain or head movement were also excluded from enrollment. Subjects who were unable to complete individual VVP-10 sessions within the allotted maximum time of 30 minutes after training were excluded from test-retest and correlation analyses.

### VVP-10 Remote Training of Patients in Their Homes

Subjects were given a mobile VR headset (Oculus Go, Facebook, Menlo Park, CA) either in-clinic (if they happened to be in clinic) or else shipped via mail. Once at home with the device, each subject was individually trained over Zoom by the investigators (ZKC, MLT) on how to take VVP-10. During the training session, the investigators helped each the subject connect the device to WiFi and discussed each step of the test (Figure 1) and the protocol for repeat testing. Patients were instructed on how the test works. Before each stimulus presentation, the test-taker performed a fixation task: they controlled a dot (the “pointer”) by moving their head, and the first step of each trial was to connect (partially overlap) this pointer with a fixation point. 0.10 seconds after successful connection, the stimulus flashed in the periphery and the subject moved the pointer towards the location of the stimulus, again by moving their head. If the subject did not see the stimulus, they could remain still until the next fixation task, resulting in a miss due to a “skipped” stimulus. Moving to the wrong location (> 30º away in polar angle) was also counted as a “miss.” This cycle of fixation task followed by stimulus task continued until all stimuli had been presented at which point the test was complete. After these instructions were given to each subject, they completed a training module within the VR testing environment during which the investigators were available online to troubleshoot. After successful completion of the training module and demonstration of an understanding of how to take the test, determined by having subjects explain the steps of the test back to the investigators, subjects were asked to complete 10 identical tests (or sessions) over 14 days. Subjects were allowed to complete up to two sessions per day, but these sessions had to take place at least 30 minutes apart to minimize fatigue from prolonged testing. At the end of the 14-day period, the investigators asked subjects to rate the levels of discomfort and fatigue they experienced from the VVP-10 test using a 5-point Likert scale (1 = no discomfort or fatigue, 5 = extremely high discomfort or fatigue). Subjects were also asked to rate the levels of discomfort and fatigue they experienced from their prior in-clinic HVF testing.

**Figure 1:**
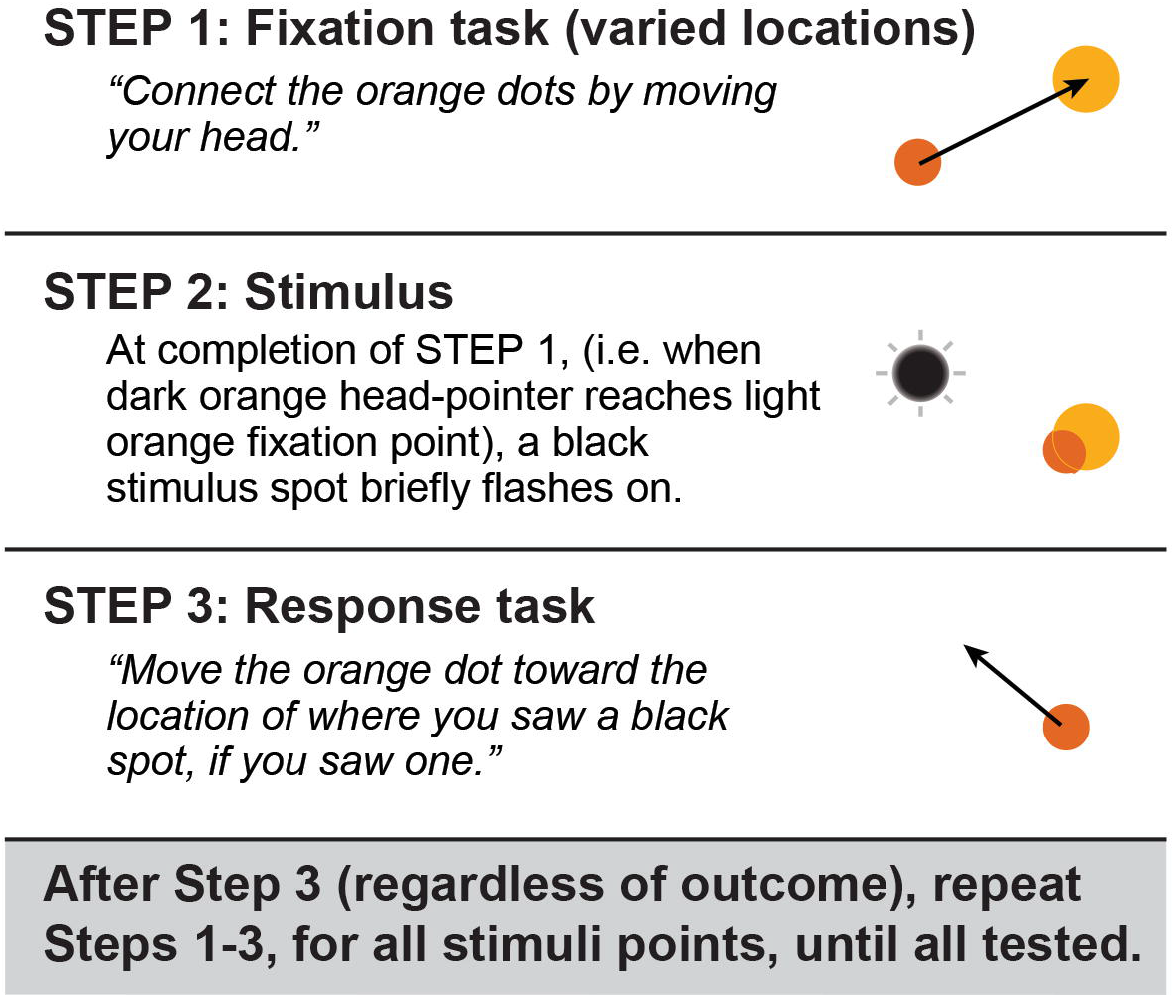
Schematic describing the steps of the test. Subjects control a pointer with their head-movement and must connect this pointer to a fixation point (fixation task). Upon connection of the head-pointer with the fixation point, a stimulus will flash in the periphery (stimulus task) and subjects must move the head-pointer in the direction of the stimulus. This cycle repeats until all points of a 24-2 grid have been tested.

### Visual Field Technical Details

The current study used VVP’s “24-2-M-FC” test, a fixed-contrast test in which all stimuli had the same suprathreshold contrast. Ten sessions of this 24-2-M-FC test comprised the VVP-10 testing strategy. Preliminary data collected using fixed-contrast stimuli demonstrated an approximately linear relationship between VVP-10 average fraction seen and HVF mean sensitivity in dB, with an average fraction seen of 50% approximately equivalent to a mean sensitivity of 15 dB. The test presented a black-on-white, blurred stimulus that was 0.43° in diameter at half-height, lasting 0.30 seconds. This stimulus was presented at 54 points on a 24-2 grid, four times per location, in a random order, and at a canonical blind spot location of (+/-14.0, -1.5 deg) seven times in each eye (total of 2 eyes x 54 locations/eye x 4 stimuli/location = 432 stimuli per test, plus 14 blind-spot stimuli). Background luminance was 25 cd/m^2^. Fraction seen was the dependent variable; the graded (shallow-slope) nature of the psychometric function causes fraction seen at a given location to vary as a function of stimulus intensity^20,21^; it likewise causes fraction seen for a fixed-intensity stimulus to vary from 0 to 1 depending on sensitivity. Because the stimuli were presented at an optical distance of ∼1.4 m (0.7 D), subjects were instructed to continue wearing any distance correction if applicable. All such subjects were able to wear their distance correcting contact lenses or glasses during testing. Progressive lenses were allowed because the test uses pattern-blurred stimuli that make it robust to small amounts of optical blur.^22^ When subjects were in clinic, their visual fields were also assessed using conventional SAP testing with the HVF 24-2 SITA-Standard test (Carl Zeiss, Meditec, Inc. Dublin, CA). These tests were not necessarily done on the same day as study enrollment, in which case data from the most recent HVF was obtained via chart review.

The fraction seen was measured at each location in each session, and average fraction seen for each eye for each session was computed. Average fraction seen was also computed for each eye across VVP-10 as a whole (across the 10 sessions). Point sensitivity and mean sensitivity (MS) for each eye in dB were the outcome measures for HVF. The cutoffs for reliability indices of HVF were set at a 15% false-positive response rate, a 30% false-negative response rate, and 30% fixation loss rate. VVP-10 does not utilize the same reliability indices and does not incorporate eye-tracking technology to determine fixation losses, but instead uses a fixation task.^23,24^ It also measures a “blind spot seen (BSS) rate,” which is the rate at which stimuli presented at the canonical blind spot, that should be missed, are seen.

### Statistical Analysis

Test-retest variability of VVP-10 was determined using the following methods. The intraclass correlation coefficient (ICC) estimate for average fraction seen, across ten VVP sessions, and its 95% confidence interval, were calculated based on a mean rating (k = 10), absolute-agreement, two-way mixed effects model. The ICC is one measure of how similar the 10 sessions were to one another. The standard error (SE) for the mean across all 10 sessions was calculated. For a given eye, the SE estimated the test-retest variability (SD) in units of the dependent variable, namely fraction seen, for the 10-session VVP-10 test as a whole.

Comparison between HVF sensitivity in dB and VVP-10 fraction seen was evaluated by calculating the Spearman correlation coefficient across an eye’s tested locations, along with its 95% confidence interval using a non-parametric bootstrap that resampled at the individual level with replacement (1,000 iterations). A separate correlation was done across eyes, using MS for HVF and average fraction seen for VVP-10. Comparison of the discomfort and fatigue that subjects experienced during VVP-10 versus HVF were calculated using the Mann-Whitney test. Significance was defined as *P* < 0.05. Data analyses were performed with R version 4.0.4 (R Foundation for Statistical Computing, Vienna, Austria).

## Results

A total of 21 unique subjects and 41 eyes were enrolled in the study. Six subjects received the headset via mail and the rest were given the headset in clinic during a regular care follow-up visit. Fifty-two percent of subjects identified as male, the rest as female, and subjects identified as either Caucasian (62%) or Asian (38%). Of the 21 subjects who were enrolled and trained remotely, one subject was considered to have failed remote training due to their inability to progress past the training module during the training session and consequent inability to complete the VVP-10 sessions. All other subjects (95%) were able to independently complete the individual sessions within the allotted time limit and successfully completed 10 sessions within 14 days after one remote training session. Figure 2 shows the grayscale maps from one subject’s 10 sessions and for the VVP-10 test as a whole, alongside their HVF grayscale maps.

**Figure 2:**
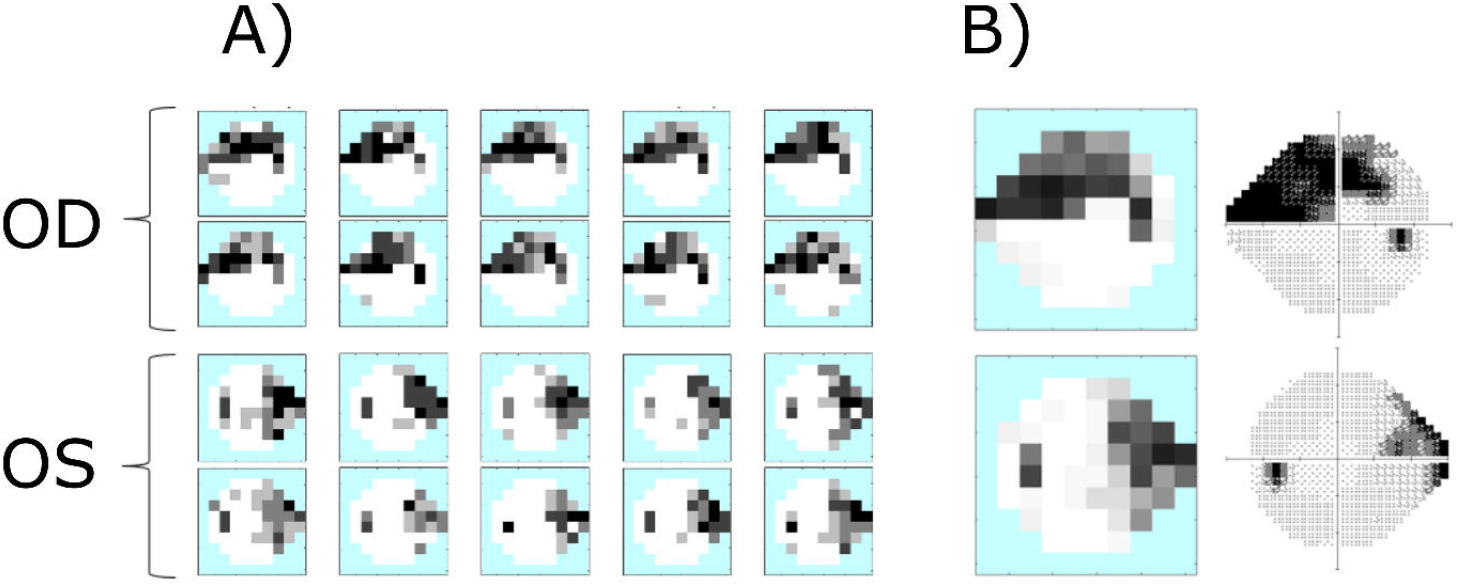
VVP-10 and HVF grayscale maps for one subject. A) Example of ten VVP-10 session results (top two rows, right eye; bottom two rows, left eye). Each location is represented by one of 5 gray levels, depending how many stimuli were seen from 0 (black) to 4 (white). B) VVP-10 grayscale map with 41 gray levels after averaging results from ten individual sessions (left column). The corresponding HVF grayscale maps for these eyes is shown in the right column. Data are from OD (top) and OS (bottom).

Of the 39 eyes from the 20 subjects who completed VVP-10, three eyes were excluded posthoc from analyses as they did not meet inclusion criteria due to the presence of concurrent ocular disease (N = 1) or BCVA worse than 20/80 (N = 2). A total of 36 eyes from 19 subjects were included in the test-retest analyses. Group characteristics of included eyes are described in Table 1. The mean time elapsed between HVF and VVP-10 testing was 32 days. On average, the response time to “seen” stimuli on VVP-10 was 0.62 seconds compared to 1.00 seconds for “missed” stimuli when the patient responded incorrectly (*P* < 0.001). The average response time for no-response “skipped” stimuli was 2.3 seconds due to the setting of the test’s time-out parameter. All eyes except one had VVP-10 blind spot seen rate < 30%.

For this set of glaucomatous eyes, the mean average fraction seen was 0.74 ± 0.21 (range 0.32 to 0.99). The intraclass correlation coefficient (ICC) of average fraction seen across the ten sessions was 0.95 (95% CI [0.92 – 0.97]) and the SE in units of fraction seen was 0.012 on average. The SE for the 10 sessions estimates the SD of their mean or the test-retest variability of VVP-10.

For comparison between HVF and VVP-10, one additional eye was excluded due to having an HVF false-positive response rate > 15% for a total of 35 eyes used in concordance analysis. Because the version of VVP-10 in this study employed a suprathreshold stimulus, it could not measure sensitivity at locations with early loss. Indeed, when correlations between HVF point sensitivity and VVP fraction seen in each eye were plotted against HVF MD (Figure 3), there was an inverse relationship between correlation and HVF MD for eyes with MD values better than -6 dB. We therefore examined the correlation between VVP-10 average fraction seen versus HVF MS for the 16 eyes that had HVF MD of -6 dB or worse. The correlation was 0.88 (95% CI [0.66, 0.99], *P* < 0.001, Figure 4A). Including all 35 eyes, the correlation between VVP-10 average fraction seen versus HVF MS was, as expected, worse at 0.68 (95% CI [0.29, 0.94]), *P <* 0.05, Figure 4B).

**Figure 3:**
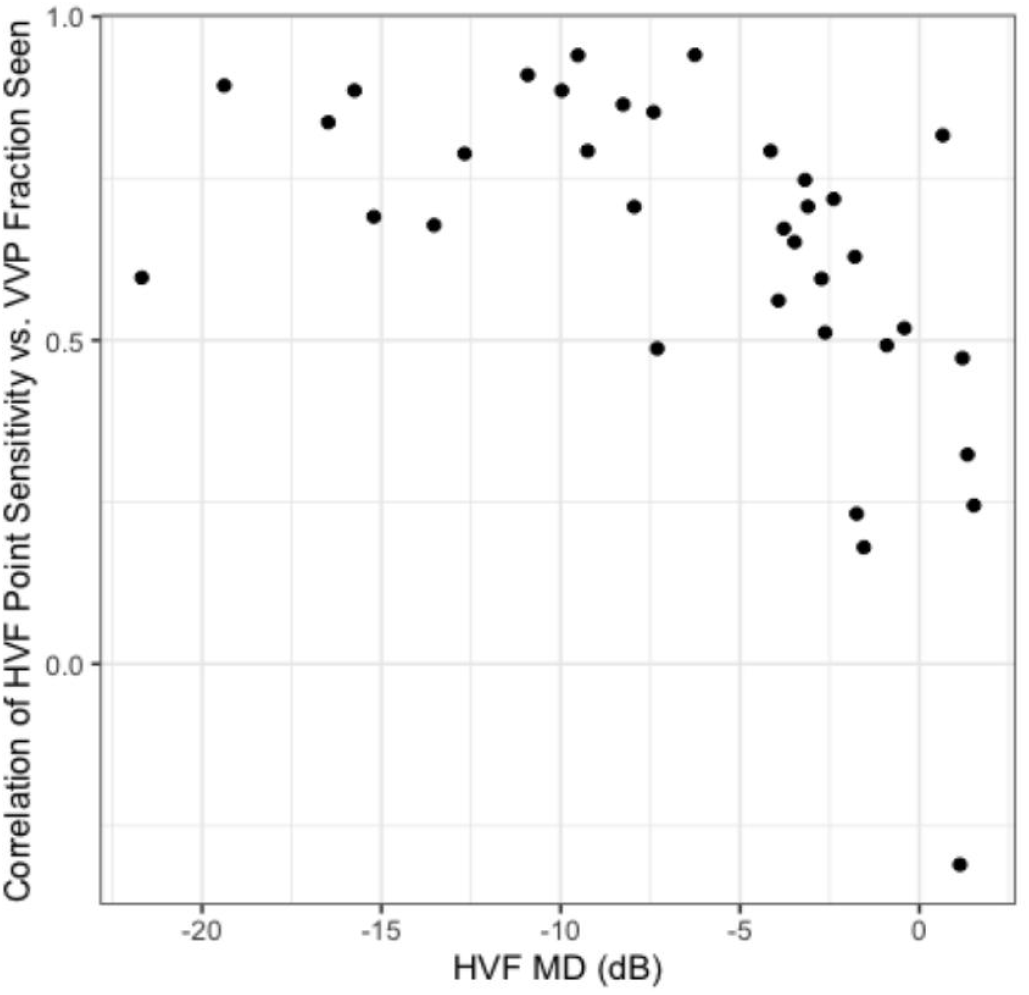
Relationship between the correlation of HVF point sensitivity (dB) versus VVP fraction seen and HVF mean deviation (dB). Each data point represents an eye.

**Figure 4:**
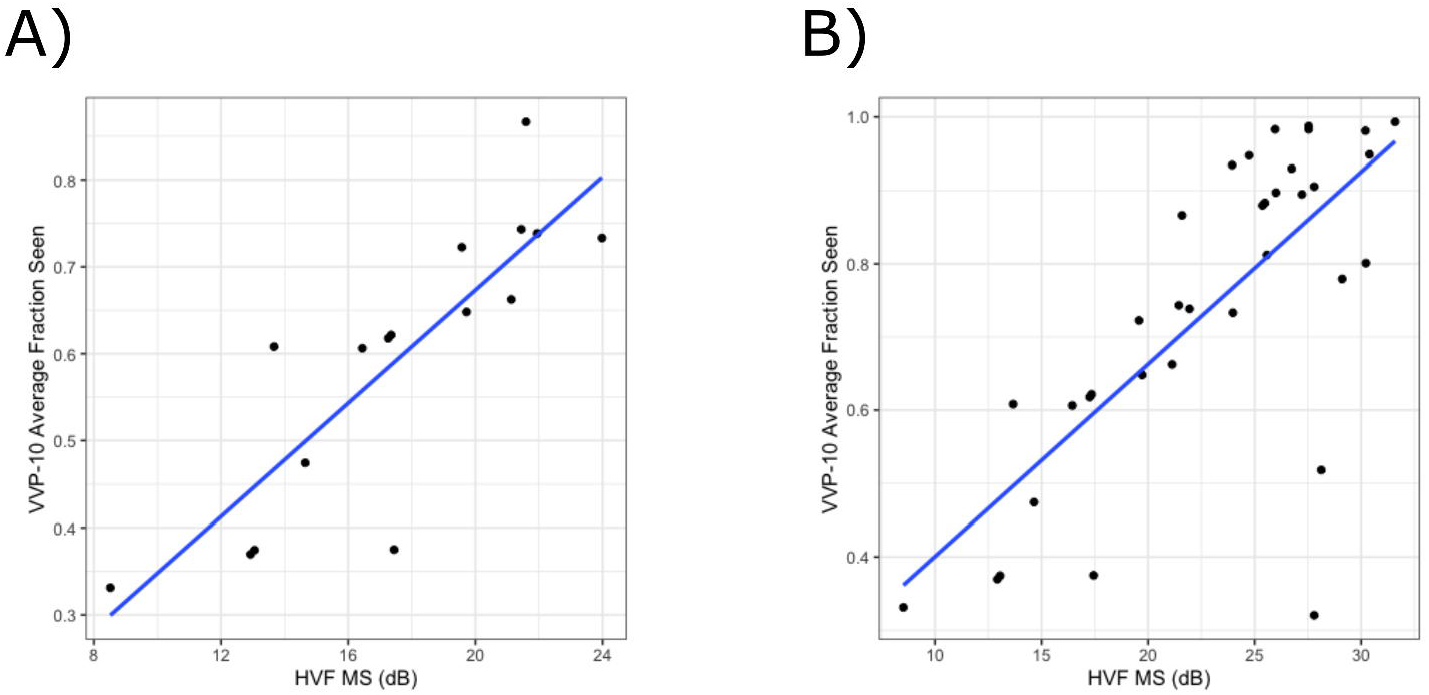
Correlation between VVP-10 average fraction seen versus HVF MS (dB) including A) only eyes with MD -6 dB or worse (16 eyes total) and B) all 35 eyes included in the correlation analysis.

When comparing one session of HVF to the ten-session VVP-10 test, patients on average felt that the tests were similarly uncomfortable (average Likert score of 2.0/5 and 1.8/5, respectively; *P* = 0.51) and similarly fatiguing (average Likert score of 2.2/5 and 1.7/5, respectively; *P* = 0.09).

## Discussion

In this study, we demonstrated the feasibility of training subjects remotely in their homes to take a new at-home VR-based visual field test, VVP-10, and the feasibility of their completing its ten sessions over a 14-day period. We estimated the test-retest variability (SD) of the mean VVP-10 test result to be 0.012, and we interpret this value as demonstrating low test-retest variability. In Figure 4A, VVP-10 average fraction seen ranged from 0.3 to 0.8. Thus, our observed test-retest variation of 0.012 was 2.4% of the range of the data. For the same eyes, HVF MS ranged from 8 to 24 dB. If it could be measured with an equivalent precision of 2.4% within this comparable range, its test-retest variability would be only 0.38 dB, which is lower than the 1.0 dB expected for glaucoma patients who are moderately reliable test takers.^25^ There is also a strong correlation between VVP-10 average fraction seen and HVF MS when subjects with mild glaucoma are excluded, which was appropriate because VVP-10 stimuli were suprathreshold.

SAP is routinely used in clinic to detect and monitor visual field defects, but many patients do not undergo the recommended frequency of testing and many dislike the testing experience. As a result, various groups have developed alternative tests that can be completed by patients at home for improved monitoring of glaucomatous changes, including Eyecatcher and Virtual Field. In studies of those tests, most patients completed tests remotely, noted the advantages of greater accessibility, and reported feeling that they were playing an active role in their healthcare.^16,26^ Patients enrolled in a study using the tablet-based Melbourne Rapid Fields test had good compliance to weekly monitoring over a 12 month period, with 32% of enrolled subjects completing the trial.^27^ To the best of our knowledge, our study is the first to demonstrate the feasibility of training subjects using video conferencing so that the entire process of training and data collection can be completed remotely.

Only one subject in our study was unable to progress past the training session, suggesting that remote training of select patients is feasible and that most patients may not need to undergo in-clinic training prior to being given a device for home testing. The reason for failure in this one subject remains unclear but likely involves some combination of challenges with the technology and a poor understanding of utilizing multi-fixation perimetry to register responses with head movement; this subject’s age at testing was 83 compared to the average age of 62.2 years. Nevertheless, the low test-retest variability of VVP-10 coupled with successful completion of the VVP-10 test series in all other subjects highlights the potential for remote training and administration of a portable visual field test. The overall correlation of 0.88 between VVP-10 average fraction seen and HVF MS in subjects with moderate to advanced glaucoma is similar to correlations obtained using other virtual reality-based head-mounted devices^28–31^ as well as tablet-based perimeters.^32,33^ Finally, although VVP-10 contained 10 sessions, subjects on average did not rate VVP-10 as more fatiguing or uncomfortable than a single HVF test. The finding that patients are amenable to taking tests more regularly has previously been described by various groups,^15,16,34,35^ but we consider it extraordinary, and an indication of our success in devising a usable test, that 95% of patients successfully completed the training and that all trained patients completed their 10 sessions without further prompting.

Strengths of this study include the remote training of subjects in their homes by two different investigators, suggesting that successful remote training does not require superior instruction by an exceptional trainer. That subjects required only one training session indicates the practical application and feasibility of remote training. We enrolled subjects who met our inclusion criteria in the order of their clinic visits and did not seek to recruit younger, potentially more technologically literate subjects.

Limitations of this study include a relatively small sample size and that all subjects identified as either Caucasian or Asian. Additionally, although we did not intentionally recruit younger subjects, the average age of our study population was 62.2 years, so our subjects may have been more comfortable using video conferencing for training than the average glaucoma patient would be. Future studies examining the utility of remote training should therefore seek to recruit older patients who may not be as familiar with video technology. VVP testing time per eye per session was also roughly double that of HVF testing. One qualitative study found that patients are willing to undergo increased duration of testing if it leads to more accurate information about their vision,^35^ but it cannot be assumed that all patients would be amenable to such prolonged testing. On the other hand, the overall effort and time to take all 10 sessions of VVP-10 at home may be similar to the time required to schedule a clinic appointment, travel to and from the clinic, and wait in the clinic.

In summary, we have demonstrated the feasibility of remote training and at-home testing, and the low test-retest variability, of VVP-10, a suprathreshold virtual reality-based visual field test. Results correlated well with in-clinic visual field testing in eyes with moderate to severe visual field loss from glaucoma. Future work should address the feasibility of remote training in older and more ethnically diverse subjects and also focus on the development of a testing strategy for longitudinal visual field monitoring. Should these studies prove VVP-10 effective for meeting those clinical needs, patients could in the future receive a testing device via mail, undergo remote training, and complete the VVP-10 test series without the burden of coming to clinic. At-risk patients who would otherwise forfeit coming to clinic in order to limit exposure to COVID-19, and those living in remote locations, could utilize VVP-10 to assess their visual function, potentially providing equivalent or complementary data to in-clinic visual field testing.

## Data Availability

All data produced in the present study are available upon reasonable request to the authors.

## Abbreviations

SAP: Static Automated Perimetry
HVF: Humphrey Visual Field
VVP: Vivid Vision Perimetry
MD: Mean Deviation
MS: Mean Sensitivity
SD-OCT: Spectral-Domain Optical Coherence Tomography
GCC: Ganglion Cell Complex
SE: Standard Error of the mean

